# 75 years’ journey of malaria publications: what and where?

**DOI:** 10.1101/2023.11.15.23298567

**Authors:** Nimita Deora, Sonalika Kar, Veena Pande, Abhinav Sinha

## Abstract

**Background:** Malaria is a life-threatening and ancient disease that has inflicted serious morbidity and mortality across the globe. The major brunt of the disease has been on African, South East Asian and South American countries. Proportionally, malaria has attracted global research priorities amongst infectious diseases and this is evident from the number of publications directly or indirectly related to malaria from across the globe, irrespective of the endemicity of the disease. However, formal and exhaustive analyses of these ‘malaria publications’ are rarely done and published.

**Methods:** We therefore did a systematic analyses of the literature published on malaria with an intent to retrieve information on what has been published on malaria, where is it published and which countries are major contributors in malaria research. The study presents malaria publications from 1945 to 2020 retrieved using three global databases: Web of Science™, Embase® and Scopus®. Exported data was examined to determine the number of publications over time, their subject areas, contributions from various countries/organizations, and top publishing journals.

**Results:** The total number of published records on malaria ranged from 90,282-1,12,698. On the basis of the number of publications, the United States, United Kingdom, France, and India were identified as the top four countries. Malaria Journal, American Journal of Tropical Medicine & Hygiene and PLoS One were the most preferred journals whereas the University of London, the London School of Hygiene and Tropical Medicine, and the University of Oxford appeared to be the top contributing organizations.

**Conclusion:** The study concluded that a disproportional contribution to malaria research was observed with non-malaria endemic countries making the largest contribution. Databases were found to be incomparable and must be standardized on multiple dimensions before making a comparison. There are still some pertinent outstanding questions that need to be addressed: why there is still a research mismatch between endemic and non-endemic regions? why a standardized and comparable information is still not available for direct and quick analyses? is less case-based research conducted as compared to in-vitro or in animal models?

## Background

Research translation (RT) or communication of the findings of one’s research to a wider audience is the most vital activity in the domain of research in any discipline. This particularly is more important when the research is related to life sciences or biomedical field wherein the effects of communicating research outputs might lead to betterment of health and/or generates evidence capable to drive the genesis or modification of an existing health policy. RT can be directed towards general public or to technical persons like other researchers, health administrators or policy makers. One of the most effective ways to translate research to fellow researchers includes publication of research in various peer-reviewed journals. The research must be published and/or documented and disseminated, otherwise, it defies the scientific purpose and ethics. It is critical to keep updated with advancements in one’s field of study and research publications are essential for the evolution of modern science, in which one scientist’s work builds on that of others (Whitesides 2004).

The timely realization of the benefits of costly medical research is a global concern that has sparked a considerable amount of legislative work around ‘translation’ (Westfall et al., 2007). Policy efforts to improve translation are in response to a large empirical literature on the challenges of transferring research from the lab to clinic (Trochim 2010; Baumbusch 2010). Both literature and policymakers presume that rapid translation of knowledge into practice is beneficial. Delays are viewed as a waste of limited resources and a potential loss of patient benefit. Timely publication of research work is much more important for diseases and health conditions which demand a quick(er) generation and transfer of new knowledge and COVID-19 is the best example which flooded the research publication arena with tons of research papers in a very small amount of time. If studies experience major publication delays or hurdles, the valuable information they provide may not reach decision-makers on time. This could be a concern for mission-oriented professions that rely on having access to the most up-to-date data in order to take effective decisions. Another such area where timely publications are of paramount importance is disease elimination and malaria (targeted for elimination by 2030) and tuberculosis (targeted for elimination by 2030) are classic examples. In any mission-driven discipline, accurate and robust access to scientific evidence is critical for effective evidence-based decision-making. Therefore, the number of scientific journal publications in a certain area per unit time is a proxy indicator of the importance of that area as is again evidenced by the sudden spurt in COVID-19-related publications.

Malaria is an ancient and life-threatening disease with publications dating as old as 3200 BC (Miller et al., 1994). Malaria is caused by nine mosquito-borne *Plasmodium* species and remains one of the most ubiquitous infectious diseases, keeping more than half the world at risk (World Malaria Report, 2022). Owing to its huge contribution in the public health burden, an estimated 247 million malaria cases were reported in 84 malaria-endemic countries (World Malaria Report, 2022). Malaria eradication remains amongst highest priorities in 86 tropical and subtropical malaria-endemic countries (Feachem et al., 2019). The estimates show that if current trends continue, malaria will considerably decrease yet remain widespread by 2050 (Feachem et al., 2019). Since malaria is targeted for global elimination in the next 7 years, it is important to analyze the malaria publications’ trend over the years, the journals which publish malaria, the subject areas within malaria that are predominant, the countries that publish the most malaria-related papers, etc. By these secondary measures, we can gauge the importance, scientific interest, political will and commitment in the field of malaria over the past years.

Systematic and scholarly information search on the topic is the need of researchers in the information-explosion era. Manually looking for information has a number of downsides that may be avoided by browsing online databases. Keeping in view the user community, there are different types of databases available which are developed and distributed through various agencies. Online databases are also made available over the internet, some are free as well as paid (they are commercial and are made available on subscription basis).

Although there are a number of databases available, each database is unique in terms of both its content and ease of use. Some database focus mainly on the content type (articles, images, videos, full-text pieces of literature or more) whereas many databases focus on only one specific subject area or disciplinary field (valuable for advanced researchers). Some databases vary in its access to content (links directly to full-text and other databases provide basic article information) whereas some are equipped with different search features. The current analyses were thus done to systematically showcase these publication trends in malaria by using 3 important databases that are said to retrieve more than 95% of available information: Web of Science™, Scopus and Embase (Bramer et al., 2017).

## Methodology

A one-time search was conducted across three major databases Web of Science™ (WOS), Embase® (EMB) and Scopus® (SCO) on May 09, 2021 using boolean operator “*Plasmodium* or Malaria”. WOS was explored through the available ‘basic search’ option with a secondary selection on ‘topic’. EMB was searched using the ‘quick search’ option where no further option was available for restricting the search term to title or abstract whereas SCO allowed using the ‘search documents’ tab for “*Plasmodium* or Malaria” in article title, abstract or keywords. No time-span filter was applied. Meeting abstracts, proceeding papers, notes, news items, corrections, reprints, biographical items, retracted publications, conference reviews, erratum, tombstone and reports published other than in English language were excluded from the final analyses.

The databases were explored during a free-trial access period provided to ICMR-National Institute of Malaria Research, New Delhi. The outcomes from 3 databases were available in two formats - entire comprehensive information (raw data) of all the published reports and pre-classified data (already analyzed data file on limited parameters). SCO did not permit the export of raw data in the trial period, hence only pre-classified data was exported from SCO whereas both raw-and pre-classified data were obtained from WOS and EMB.

Exported data were analyzed to address the number of publications over time, their subject areas, contribution from different countries/organizations and names of publishing journals. Because subject areas were not uniform across the databases, the subject areas were re-grouped into broader categories in order to make the data presentable and comparable across the databases. The top-ten ranked countries (Figure 4), organizations (Figure 9 A-B), journals (Figure 5) and subject areas (Figure 8 A-B) were then identified based on the number of publications made in each of these parameters using pre-classified data.

Apart from identifying top ranked countries (based on the number of publications), contribution of countries was reanalyzed based on their respective population. The total population of the countries in 2021 (https://datacommons.org/place) was used as a denominator for calculating publications per million population using the two databases SCO and WOS. The top 10 countries with highest number of publications in malaria and top 10 countries with highest publications per million populations were listed out from the above data as shown in Figure 3 (A-D). To identify all unique journals that published malaria over time, journal details from all EMB, SCO and WOS databases were merged, and the total number of journals was determined after removing duplications.

For determining the number of malaria publications every year from the top ten journals, the raw data from WOS was only used because it offered the longest historical data (since 1945). The raw data obtained from WOS was cleaned and sorted year-wise for each of the top-ten journals to calculate the year-wise number of publications of that particular journal (Figure 6 A-B). Furthermore, to ascertain the trend of malaria publications with malaria burden, the estimated number of malaria cases from the World Malaria Report 2021 were recorded and plotted against the year-wise malaria publications from top ten journals (Figure 7). Only WOS was used for this analysis because this was the only database which contained data from 1945.

## Results

The total number of published literature (and temporal coverage in years) on malaria retrieved from WOS, EMB and SCO were 90,282 (1945-2020; 75 years), 1,12,698 (1971-2020; 49 years) and 1,12,594 (1960-2020; 60 years) respectively (Figure 1).

**Figure 1:**
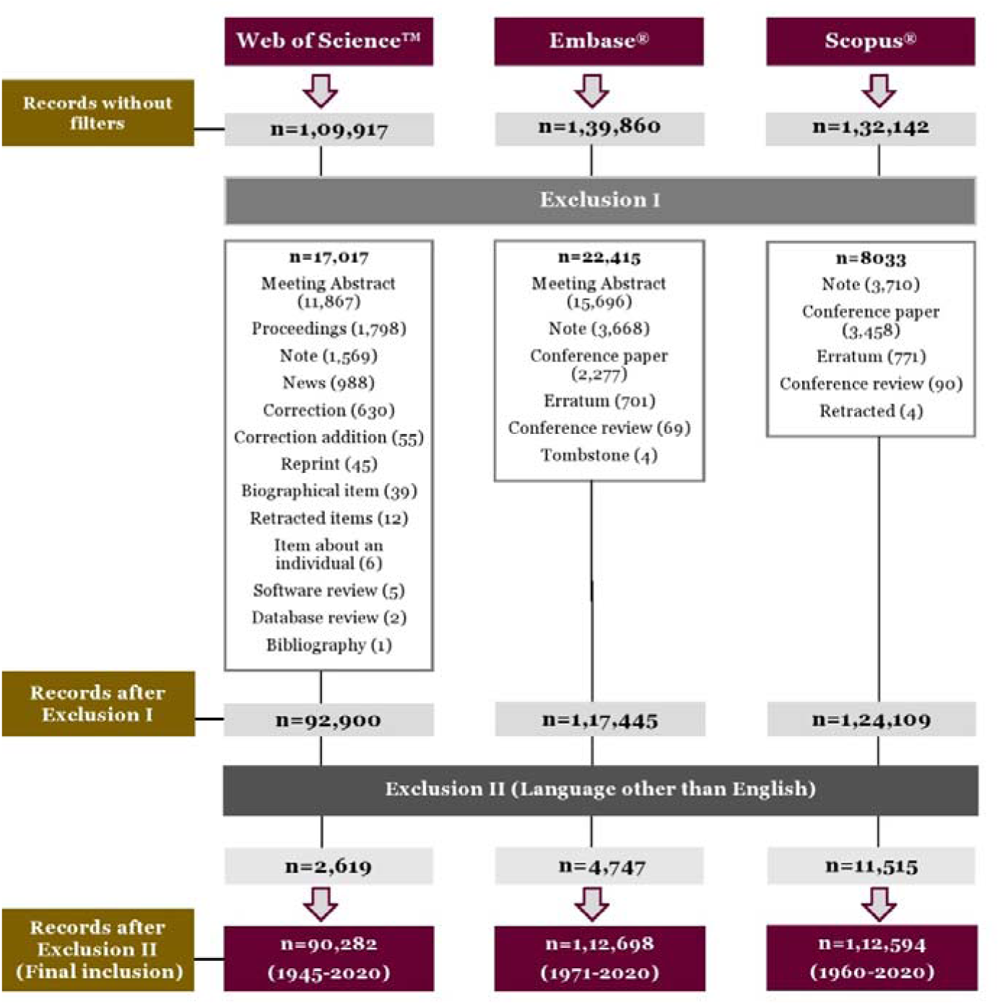
Search strategy for data extraction from Web of Science™, Scopus^®^ and Embase^®^.

All databases were scrutinized vigorously to bring forth the available underneath information that may significantly enhance our understanding towards publication trends in malaria. Records indicate that the first documents were published in 1945 (obtained from WOS). The other two search engines, SCO and EMB, provided records of malaria-related documents that were initially published in 1960 and 1971, respectively (Figure 2). Since then, researchers’ interest in the field of malaria has grown as observed by number of published documents overtime. The highest number of annual publications (n=5,517) were recorded in 2014, followed by n=5,509 in 2020 (EMB).

**Figure 2:**
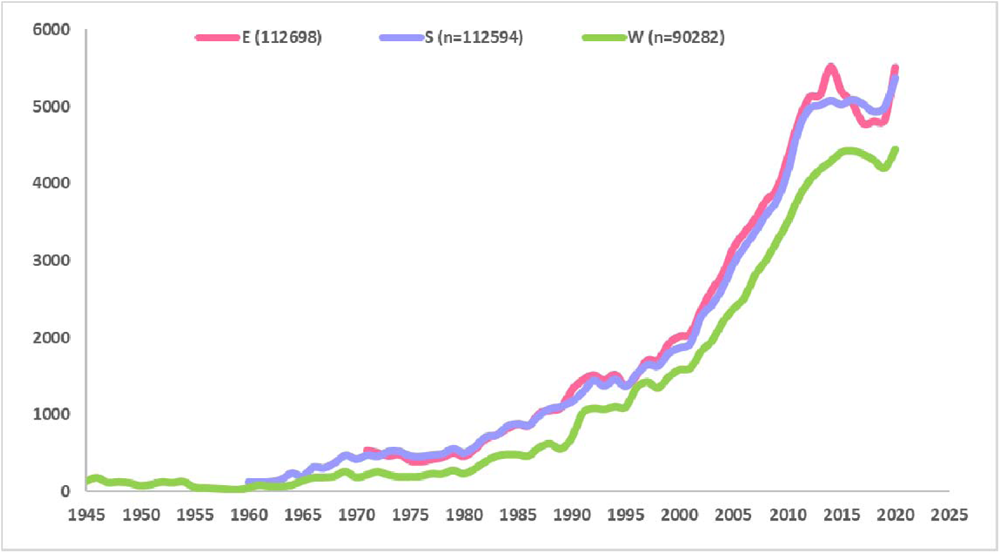
Number of annual malaria publications obtained from three different databases. The graph displays the evolution of the number of annual publications on malaria over time obtained from three databases Embase®(E), Scopus® (S) and Web of Science™ (W). Here, x-axis and y-axis display year and number of publications, respectively.

Between 1945 and 2020, of all the malaria publications in the WOS (n=1,50,239), 20% (n=29,313) has been published by the United States (US), followed by UK (n=15,368; 10%), and France (n=6,889; 5%) (Figure 3A). Despite India being one of the most affected countries by malaria, it contributed only 4% (n=6,276) of publications, which is 5X less as compared to the US. Similarly, SCO (n=1,80,486) published 18% (n=33,191) research on malaria done by the US, with 11% (n=18,961) contribution from the UK and India made 5% (n=9,542) of the total publications during 1960-2020 (Figure 3B). Other developed and underdeveloped countries published between 0.01 & 2% in both the databases. However, when the publication data was standardized by the country-specific, French Guiana, Switzerland, The Gambia, Scotland/UK and Gabon were the top 5 countries publishing on Malaria as per WOS and SCO but their ranks differed between the databases as shown in Figure 3 (A-D). It is to be noted that Switzerland, England/UK and Australia were the only 3 countries that retained their slots in the top 10 countries publishing the most in malaria in both WOS and SCO.

**Figure 3.**
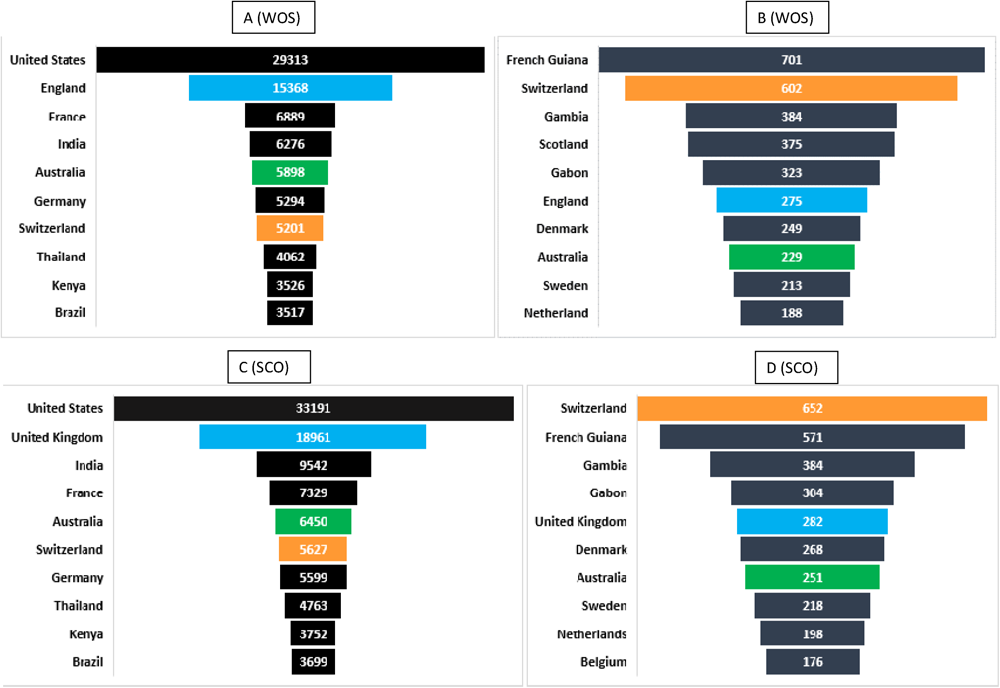
(A-D): Top ten countries on the basis of number of publications (A & C) and publications per million populations (B & D) Figures A and C were created by plotting the number of publications from a specific country from 1945 to 2020, as obtained from WOS and SCO, respectively. Figures B and D show the countries based on their publications per million populations as determined by WOS and SCO, respectively. The total population of the countries in 2021 was used as a denominator in t calculation to calculate publications per million population of a specific country. Countries that consistently appeared in the top ranked list were colored the same in both figures (A-B & emphasize their position.

Here, the denominator (total number of published records) was taken from pre-classified data of WOS and SCO exported during data-curation. Therefore, the denominator varies depending on the parameters examined. The denominator in country-wise pre-classified data retrieved from various databases has increased from that obtained from year-wise data because each record was counted once in year-wise analysis but records may have been counted more than twice in country-wise analysis if they have affiliations from >1 countries. Each parameter, similarly, has its own denominator. EMB did provide such country-wise information in the pre-classified data and hence, it was not included in the above analysis.

The number of publications obtained from the SCO (n=1,80,486) was used to rank countries from one to ten in Figure 4. Similar results were obtained from WOS except for India and France. As per WOS data (n=1,50,239), India was ranked fourth (n=6,276; 4%) and France (n=6,889; 5%) was ranked third. All remaining countries maintained the same position as SCO. It should be noted that, despite being a malaria non-endemic country, the US is the largest contributor with 18-20% of the total publications, with the remaining individual countries falling below 11%. The top ten countries contribute between 2 and 20% each. Based on data obtained from various databases, the least contributing countries were also identified (shown in supplementary data) which contribute to <1%.

**Figure 4:**
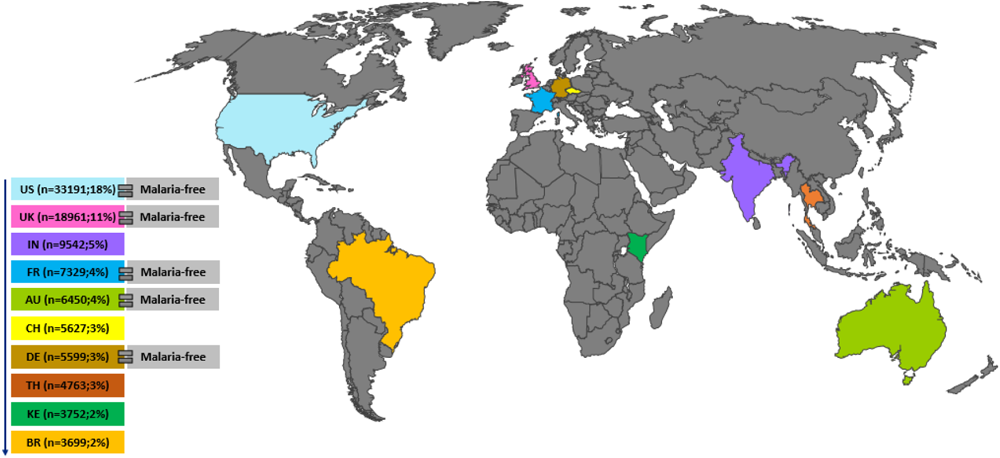
Top ten countries with the highest number of malaria related publications. Here, the highest contributing countries have been shown in different colors whereas countries with a negligible contribution to malaria publications are represented by the grey shaded area. In the left panel, colored boxes represent top ten countries (on the basis of data obtained from Scopus®) and are in descending order of their contribution to malaria publication between 1960-2020. Here, ‘n’ represents number of published records on malaria. US: United States, UK: United Kingdom, IN: India, FR: France, AU: Australia, CH: Switzerland, DE: Germany, TH: Thailand, KE: Kenya, BR: Brazil

**Figure 5:**
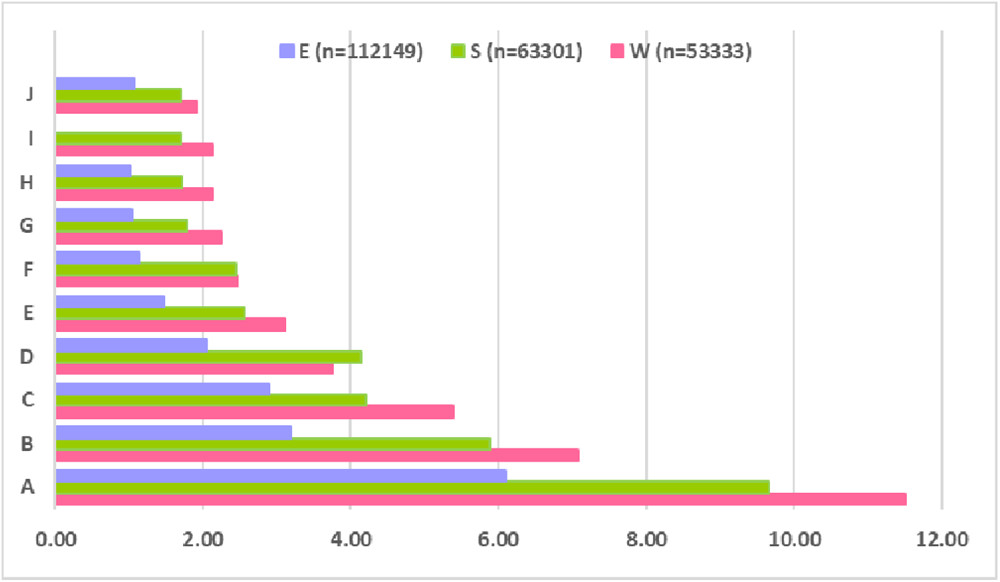
Journal-wise analysis of malaria related published records over the years. The figure represents the percentage contribution of the top 10 journals ranked from one (highest; A) to ten (lowest: J) in which various malaria related records were published during 1945-2020. Here, x-axis represents percentage of published records and y-axis represents journals coded by alphabets (A-J). Records from Embase® (E), Scopus® (S) and Web of Science™ (W) have been shown in blue, green, and pink respectively. A: Malaria journal; B: American journal of tropical medicine and hygiene; C: PLoS One; D: Transactions of the royal society of tropical medicine and hygiene; E: Molecular and biochemical parasitology; F: The Lancet; G: Infection and immunity; H: Antimicrobial agents and chemotherapy; I: Experimental parasitology; J: Journal of infectious diseases

**Figure 6.**
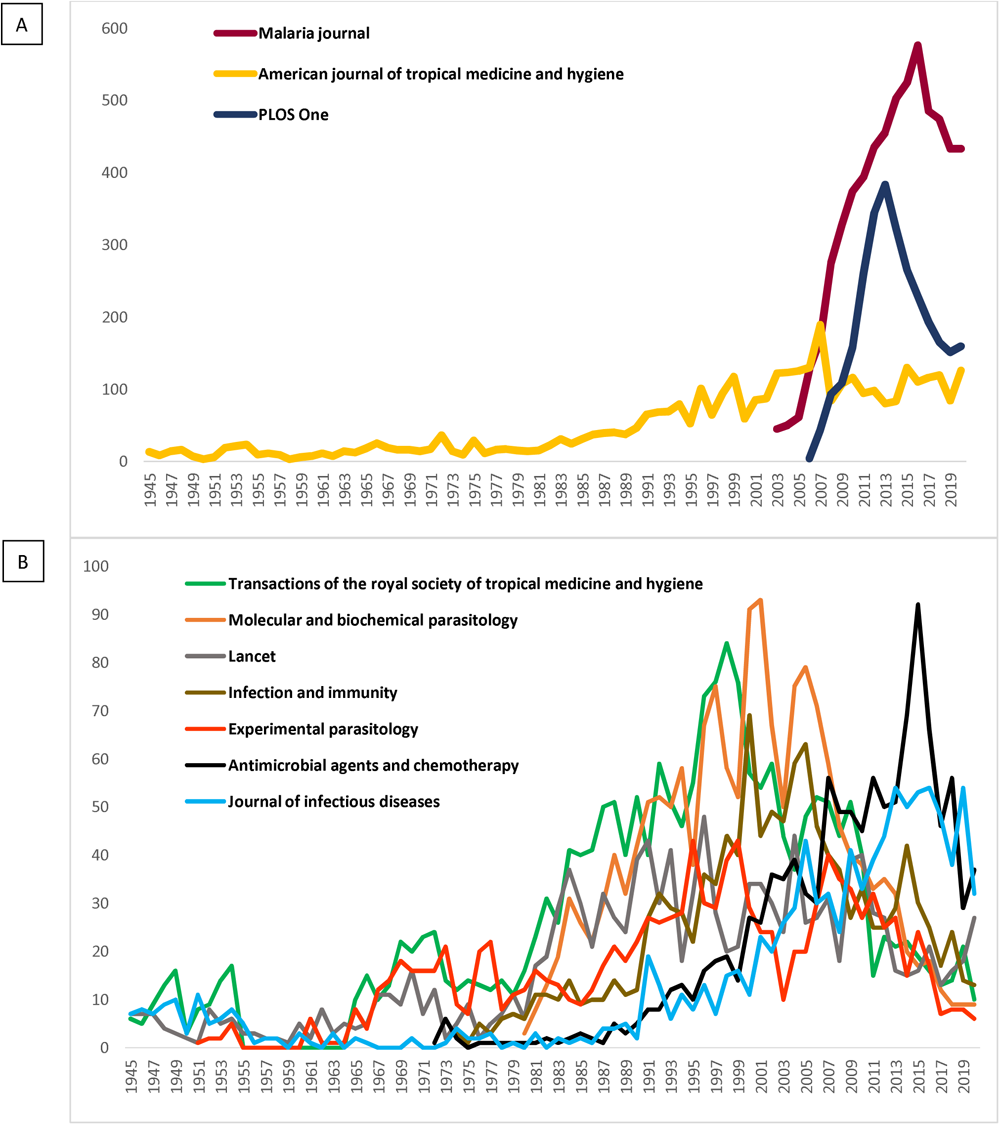
(A-B): Performance of the most active journals ranked from one to ten (on the basis of number of publications) between 1945-2020. Figure 6A depicted the top three journals Malaria, ASTMH, and PLOS One, while Figure 6B depicted the remaining seven journals. The Y axis represents the number of publications, while the X axis represents the year of publication. It is to be noted that the launch year of all journals was not the same.

**Figure 7:**
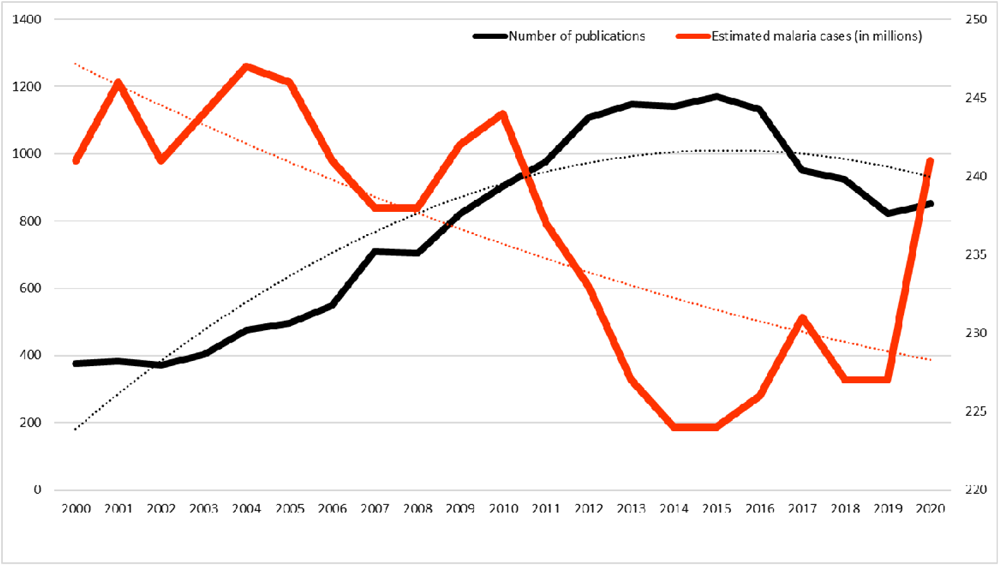
Correlation between the number of publications and the number of estimated malaria cases worldwide from 2000 to 2020. The secondary axis used to plot the global estimated malaria cases here represents numbers in millions while the Y-axis represents the total number of publications from the top 10 journals. Their respective colored dotted lines show the historical trend of the data.

All the retrieved records from the three databases found to be published in 1811 different journals. When it comes to the choice of publication platform, the Malaria Journal is the most preferred platform of researchers around the globe to showcase their findings as seen from the trend of publications in various journals indexed by WOS (n=6,133/53,333; 11.5%), SCO (n=6,123/63,301; 9.7%) and EMB (n=6,835/1,12,149; 6.1%). The top five preferred journals by researchers included Malaria Journal, American Journal of Tropical Medicine And Hygiene, PLoS One, Transactions Of The Royal Society Of Tropical Medicine And Hygiene and Molecular And Biochemical Parasitology. The number of records published in the top ten journals in WOS, SCO and EMB was 22,256 (42%), 22,675 (36%) and 23,545 (21%), respectively. It was found in WOS that almost half of the total records were published in just ten journals (more than one-third in SCO and one-fifth in EMB).

It is clear from figure 8 that more than one-thirds of the total records were related to medicine (WOS 45%; SCO 37%). Here, it is to be noted that in order to get uniform data from two different databases, subjects were grouped into one category if they were initially found scattered across sub-categories. Other relevant subjects under which malaria related records were published were found to be immunology and microbiology, biochemistry, genetics and molecular biology, agricultural and biological sciences, pharmacology, toxicology, and pharmaceutics and public, environmental and occupational health. Remaining less common subjects were chemistry, multidisciplinary sciences, veterinary sciences, environmental science and social sciences under which less than 5-6% of malaria related records were published between 1945-2020. Further, such pre-classified data was not available in EMB and hence was not analyzed for this particular category.

**Figure 8:**
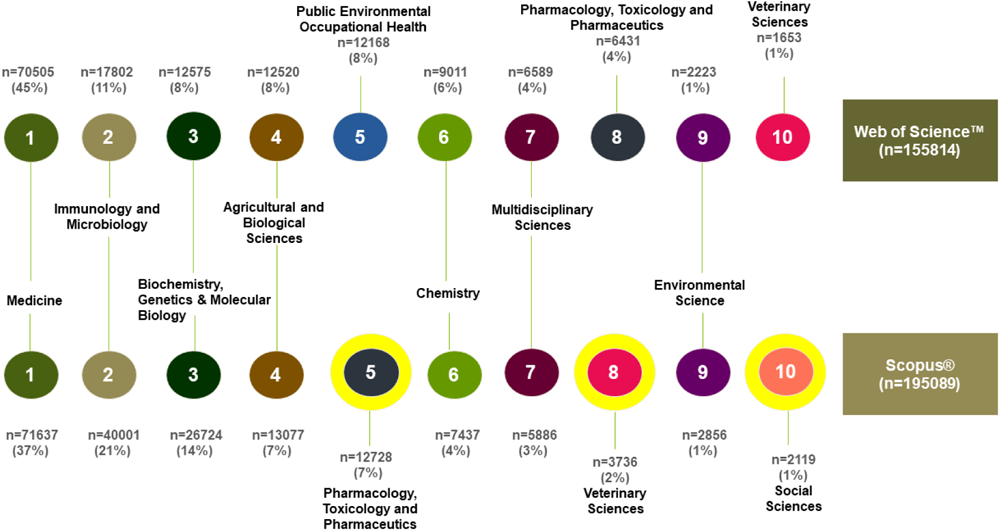
Subject-wise analysis of malaria related published records over the years. The figure depicts the top ten subjects (as determined by the Web of Science™ & Scopus® databases) under which malaria-related literature were published over time. A specific subject has been highlighted with a specific colour in both databases. Subjects have been listed from one to ten in descending order based on the number of publications. Where a difference in a sequence was observed in both databases, subjects have been labelled with a secondary yellow circle and marked separately with a particular subject name. Here, ‘n’ represents number of records published.

Based on WOS (1945-2020), 1,16,365 records on malaria were found to be published from 100 organizations around the world after analyzing the research output of various organizations and groups whereas 1,12,681 records were found in SCO (1960-2020) database, published from 162 organizations worldwide. The contributions from various organizations have been widely dispersed. However, the top contributing organizations have been depicted in figure 9 (A-B), which contributed between 2% and 5% of total publications.

**Figure 9.**
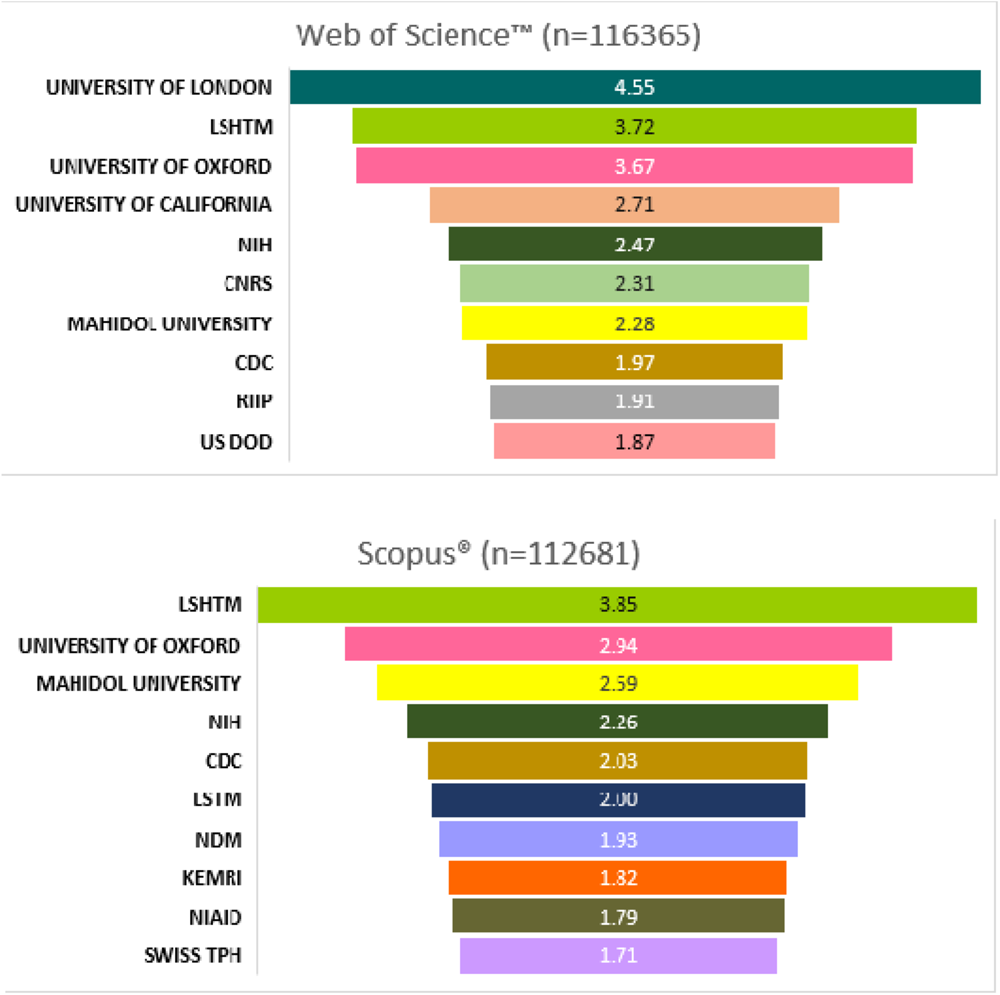
(A-B): Highest contributing organizations to the global malaria research. This figure depicts the most active scientific organizations dedicated towards malaria research based on the data obtained from Web of Science™ (9A) & Scopus® (9B). Here, top ten contributing organizations have been arranged in descending order of their contribution towards malaria research and the number represent percentage of their publications out of total published records. LSHTM: London School of Hygiene Tropical Medicine, NIH: National Institute of Health, CDC: Centre For Disease Control Prevention, LSTM: Liverpool School Of Tropical Medicine, NDM: Nuffield Department of Medicine, KEMRI: Kenya Medical Research Institute, NIAID: National Institute of Allergy and Infectious Diseases, SWISS TPH: Swiss Tropical and Public Health Institute, CNRS: The French National Centre for Scientific Research, RIIP: Réseau Institut Pasteur International, US DOD: United States Department of Defense

As evident from WOS (Figure 9A), the University of London published ∼5,300 (5%) records which is the highest amongst all other organizations. London School of Hygiene & Tropical Medicine (LSHTM; n=4,333) and University of Oxford (UoO; n=4,276) contributed ∼4% of total publications, each. Other organizations from top ten ranked list contributed less than 3% each. One of the largest fundamental science agencies in Europe, the French National Centre for Scientific Research, contributed ∼2% (n=2,685) publications on malaria. Mahidol University (n=2,653) and the Centers for Disease Control and prevention (CDC; n=2,291) also contributed ∼2% each. When the Scopus® database was examined, sequential variations were observed. The top three contributing organizations were the LSHTM with 4,334 publications (4%), UoO with 3,310 publications (3%), and Mahidol University, with 2,917 publications (3%). Furthermore, the National Institutes of Health, the CDC, and the Liverpool School of Tropical Medicine published 2543, 2292, and 2257 records, respectively with a contribution of ∼2% each. Other organizations from top-ten list contributed below 2% (Figure 9B).

## Limitations

Unlike pre-classified data, which had separate datasheets for each parameter alongside the number of publications, the raw data had all of the information in a single datasheet, including the year of publication, affiliation of authors, country, journal etc. The sole drawback of pre-classified data was that we could never correlate one statistic with another (for example, year-wise performance of a country) because it provides pre-analyzed data individually for each parameter. Therefore, in order to examine two parameters at once, we utilized raw data. Subject-wise classification: overlap due to multi-disciplinarity and no rigid boundaries between scientific disciplines, for example, medicine and immunology or pathology…affiliation for country level and/or institutional level, university of London and LSHTM!

## Discussion

The focus of this research is to provide an overview of global research publications in the field of malaria with an aim to quantify the malaria-related publications in terms of various metrics by retrieving respective literatures from three major scientific databases Web of ® ® Science™, Scopus and Embase. The bibliometric analysis method, which has lately become widely employed in numerous health themes, has been used to demonstrate the study pattern and research activity on a certain topic (Sweileh et al., 2017; Sweileh et al., 2018). Furthermore, for future funding and planning, bibliometric analysis is an appropriate tool for identifying key research themes, active researchers, and research organizations. Distinguishing proof and assessment of exceptionally referred to articles on a particular topic might be important to achieve optimized research allocation, reorientation of research support, rationalize research organizations, restrict research in particular fields or boost research productivity. The quantum and trend of publications in a specific field also serves as proxy indicators of political will, funding and scientific interest.

Although a few bibliometric studies have been performed on research related to malaria, ours is a first analysis of this kind as its targets global malaria bibliometric research through three different databases. Previously published bibliometric malaria-related research was either restricted to a particular geography (Latin America -Munoz-Urbano et al., 2014; China – Fu et al., 2015, Du et al., 2021; India – Singh and Mahanty, 2019, Gupta and Bala, 2011; Malawi – Mwendera et al., 2017; Portugal – Ferreira and Teixeira, 2019; Central African Republic - Nzoumbou-Boko et al., 2022), a particular period or a specific context related to malaria (artemisinin – Dong et al., 2022, Xu et al., 2018; Yao et al., 2012; malaria vaccines – Garg et al., 2009; malaria in pregnancy - van Eijk et al., 2012; malaria vector resistance – Sweileh et al., 2016; anti-malarial drug resistance - Sweileh et al., 2017; citations - Ghamgosar, Zarghani, and Nemati-Anaraki, 2021, Kolle, Vijayashree, and Shankarappa, 2017) or to a wider term such as mosquito-borne disease (Ong, Pauzi, and Gan, 2022) or parasitology (Ellis et al., 2020) or even COVID-19 (Mala et al., 2022). We could find only one article that analyzed global research on malaria (Garrido-Cardenas et al., 2019) but this article was focused on *P. vivax* and only from the Scopus database.

The three databases used in this study are not only have a wide coverage of articles but also have differential and complementary features. It has already been established that a reasonably good recall or coverage from a database search is not linearly associated with the number of databases searched but depends on the optimal use of selected databases (Bramer et al., 2017). Scopus, launched in 2004, offers about 20% more coverage and covers a wider journal range than Web of Science which initially started in 1963 but Web of Science covers older publications as its search period goes as back as 1900 as compared to Scopus (1966 to present) (Falagas et al., 2008; Bar-Ilan, 2018) and therefore the combination of Scopus and Web of Science offer the widest and oldest publication records in a scientific field. As Embase has additional focus and relevance to clinical sciences, its addition to Scopus and Web of Science adds more depth to clinical literature search (Mann et al., 2016; Bramer et al., 2017). Although it is suggested that a combination of Embase, PubMed, Web of Science and Google Scholar offer a near-100% overall recall (Bramer et al., 2017), we replaced PubMed and Google Scholar with Scopus as the latter offered a similar or wider journal collection and the former two did not offer pre-analysed data output. However, Embase, Scopus and Web of Science are not freely available and need a paid subscription and therefore the analyses were dependent on the information that was available during the limited-period free-trial access that was offered to our Institute in 2020. We noted a near-identical rising trend of global malaria publications with a slight dip between 2015 and 2020 in all the three databases. The rising trend might indicate a constant researchers’ interest, opportunities, funding and need for malaria research globally whereas a dip may indicate lack of commitment and complacency due to declining malaria cases around the world. This trend is more conspicuous in Figure 7 where it can be reasonably debated that a sharp decrease in malaria cases between 2010 and 2014 might have translated into a decline in funding for malaria research and therefore leading to a decrease in the number of publications between 2015 and 2020 as noted in this analyses as has been reported by multiple authors in different areas (Hussinger & Carvalho, 2022; Sattari et al., 2022; Jacob and Lefgren, 2011). However, a spurt in the number of malaria cases globally from 2015 and more recently after the COVID-19 (World Malaria Report, 2021, World Malaria Report, 2022), a concomitant response from all stakeholders to enhance commitment in malaria research funding and interest should drive up the annual number of malaria publications past the 2012-2015 levels.

In terms of the top 10 countries publishing most in malaria standardized by their respective population, only The Gambia and Gabon belonged to the malaria high-endemic regions. Nigeria, the Democratic Republic of Congo and Uganda, the top 3 countries contributing the highest to malaria cases in 2020 (World Malaria Report, 2021) were ranked as 69 & 59, 87 & 100, and 43 & 40, respectively in WOS & SCO in the total number of malaria publications across the period. Similarly, India, which contributes the highest number of cases in South-East Asia, was ranked 84 & 80, respectively in WOS and SCO. Although the manner in which a country’s contribution is counted in both the databases may differ, it is evident that malaria endemic countries do not publish much on malaria as compared to the countries with minimal or no malaria! Since Malaria Journal is the only journal which is 100% dedicated for malaria-related publications, it is no doubt the topmost journal in terms of the number of publications as reported in the three databases analyzed.

## Conclusion

This research concludes a disproportional contribution of malaria research and publications between endemic and non-endemic countries with many times higher number of publications from non-malaria endemic countries. Literature databases are not straightaway comparable with significant overlaps and exclusiveness and need to be standardized on multiple aspects before a comparison is made.

### List of abbreviations

RT: Research Translation
EMB: Embase
SCO: Scopus
WOS: Web of Science
LSHTM: London School of Hygiene & Tropical Medicine
CDC: Centers for Disease Control and prevention
UoO: University of Oxford

## Declarations

### Ethics approval and consent to participate

Not applicable

### Consent for publication

Not applicable

### Availability of data and material

Available on reasonable request

## Authors’ contributions

AS and VP conceived the idea. ND and SK conducted the systematic review and acquired the data. ND drew the figures. ND, SK, VP and AS analyzed and interpreted the data. ND wrote the first draft. ND, SK, VP and AS reviewed the draft and provided critical insights. All authors agreed to the final version of the submitted manuscript.

## Data Availability

All data produced in the present study are available upon reasonable request to the authors

## Acknowledgement

ND was supported by WOS-A scheme, Department of Science & Technology and Director ICMR-NIMR is acknowledged for providing infrastructural facilities.

